# Leveraging intelligent optimization for automated, cardiac-sparing accelerated partial breast treatment planning

**DOI:** 10.1101/2022.12.28.22284011

**Authors:** Joel Pogue, Carlos Cardenas, Yanan Cao, Richard Popple, Michael Soike, Drexel Boggs, Dennis Stanley, Joseph Harms

**Author notes:** **Correspondence:** J. A. Pogue.

## Abstract

**Background:** Accelerated partial breast irradiation (APBI) yields similar rates of recurrence and cosmetic outcomes as compared to whole breast radiation therapy (RT) when patients and treatment techniques are appropriately selected. APBI combined with stereotactic body radiation therapy (SBRT) is a promising technique for precisely delivering high levels of radiation while avoiding uninvolved breast tissue. Here we investigate the feasibility of automatically generating high quality APBI plans in the Ethos adaptive workspace with a specific emphasis on sparing the heart.

**Methods:** Nine patients (10 target volumes) were utilized to iteratively tune an Ethos APBI planning template for automatic plan generation. Twenty patients previously treated on a TrueBeam Edge accelerator were then automatically replanned using this template without manual intervention or reoptimization. The unbiased validation cohort Ethos plans were benchmarked via adherence to planning objectives, a comparison of DVH and quality indices against the clinical Edge plans, and qualitative reviews by two board-certified radiation oncologists.

**Results:** 85% (17/20) of automated validation cohort plans met all planning objectives; three plans did not achieve the contralateral lung V150cGy objective, but all other objectives were achieved. Compared to the Eclipse generated plans, the proposed Ethos template generated plans with greater evaluation planning target volume (PTV_Eval) V100% coverage (*p* = 0.01), significantly decreased heart V1500cGy (*p* < 0.001), and increased contralateral breast V500cGy, skin D0.01cc, and RTOG conformity index (*p* = 0.03, *p* = 0.03, and *p* = 0.01, respectively). However, only the reduction in heart dose was significant after correcting for multiple testing. Physicist-selected plans were deemed clinically acceptable without modification for 75% and 90% of plans by physicians A and B, respectively. Physicians A and B scored at least one automatically generated plan as clinically acceptable for 100% and 95% of planning intents, respectively.

**Conclusions:** Standard left- and right-sided planning templates automatically generated APBI plans of comparable quality to manually generated plans treated on a stereotactic linear accelerator, with a significant reduction in heart dose compared to Eclipse generated plans. The methods presented in this work elucidate an approach for generating automated, cardiac-sparing APBI treatment plans for daily adaptive RT with high efficiency.

## 1 Introduction

The incidence rate of early stage breast cancer is steadily increasing due to improved detection and screening strategies [1]. Equivalent overall survival rates of lumpectomy followed by external beam radiation therapy (RT) compared to mastectomy have been shown [2], and post-lumpectomy pathologic analysis by Vicini et al. demonstrated that residual disease occurred within 1cm of the lumpectomy cavity for more than 90% of patients [3]. Until recently, external beam accelerated partial breast irradiation (APBI) has been less preferred to brachytherapy APBI due to the large planning target volume (PTV) margins necessary to account for set-up uncertainty, resulting in increased healthy tissue exposure and inferior cosmetic outcomes relative to whole breast RT [4].

However, technical improvements in patient immobilization, imaging, and dosimetry have more recently piqued interest in stereotactic body radiation therapy (SBRT), which allows for reduced margins and steeper dose fall-off outside of the target.

To that end, Vermeulen et al. observed no toxicities ≥ grade 3 for 46 stage 1 patients receiving supine SBRT treatment with a 2mm PTV expansion [5; 6]. Additionally, Timmerman et al. published methods and cosmetic outcomes for a 75 patient, five arm dose-escalation SBRT trial in which high rates of good or excellent cosmesis were achieved [7; 8]. Livi et al. demonstrated that compared to conventionally fractionated (5000cGy in 25 fractions) breast treatment, intensity modulated radiation therapy (IMRT) based PBI resulted in significantly fewer short and long term toxicities and improved cosmetic satisfaction compared to whole breast RT using 1cm PTV margins [9]. Based on these findings, our institution initiated the UAB RAD 1802 trial (Pilot Trial of LINAC Based Stereotactic Body Radiotherapy for Early Stage Breast Cancer Patients Eligible for Post-Operative Accelerated Partial Breast Irradiation (APBI); clinicaltrials.gov identifier NCT03643861). The purpose was to combine the SBRT techniques and accelerated fractionation schemes, which were previously exclusively utilized on the Cyberknife platform, with the IMRT capabilities of a traditional linear accelerator. Methods and preliminary findings for the first 23 patients (16 prone, 7 supine) have since been published [10].

While novel platforms such as RapidPlan and HyperArc (Varian Medical Systems) have provided a means of automating planning processes [11; 12], many institutions still heavily rely on iterative, manual planning [13; 14]. Developing alternatives to manual planning would be ideal as the time required to train personnel and manually generate high-quality treatment plans remains costly [15]. Furthermore, planning skill varies greatly by planner and site [16; 17], manual plan constraints and optimization structures are often inconsistent, and time limitations greatly impact the quality of manual plans. Thus, the aim of automation is to increase plan consistency, reduce planning time, and maintain or improve plan quality. Popular forms of automation include, but are not limited to, knowledge-based planning (KBP) [18; 19; 20; 21; 22], multi-criteria optimization (MCO) [23; 24], and template-based planning [25; 26; 27; 28]. The Ethos (Varian Medical Systems, Palo Alto, CA) treatment planning system (TPS) utilizes an Intelligent Optimization Engine (IOE) that mimics the way a skilled planner controls optimization priorities and structures [29]; it automatically generates multiple plans from pre-defined beam geometries, each optimized according to an identical optimization template, or intent. Furthermore, the Ethos platform allows for RT plan adaption based on daily cone beam CT (CBCT) anatomy [30; 31], the benefits of which are promising when treating potentially mobile targets through pendulous anatomy.

Significant effort has been dedicated to sparing the heart in lung RT due to high levels of proximal dose [32; 33], but it has also been observed that breast RT induces cardiac toxicity linearly with no apparent dose threshold. Increased risk in major coronary events between 7.4% and 19% per additional Gy of mean heart dose have been reported for breast RT [34; 35; 36]. Based on these findings, the goal of this work is to illustrate the feasibility of automatically generating high-quality APBI treatment plans in the Ethos adaptive workspace with a specific emphasis on cardiac-sparing; furthermore, these plans are benchmarked against clinically approved, manual plans treated on a TrueBeam Edge linear accelerator. Adherence to RAD 1802 planning objectives, a comparison of manual and automated dose volume histograms (DVH) and quality indices, and qualitative review by multiple board-certified radiation oncologists will each be utilized to elucidate Ethos automated plan quality.

## 2 Materials and Methods

### 2.1 Cohort Description

29 patients (30 plans due to one patient with bilateral disease) previously receiving supine APBI treatment for early stage breast cancer (stages 0-2) at our institution between 2019 and 2022 were utilized in this Institutional Review Board (IRB-1207033005) approved study. Seven patients met RAD 1802 inclusion criteria and were simulated and contoured according to trial protocol. Inclusion criteria consisted of age ≥ 50, estrogen receptor (ER) positive, and negative margins of at least 2mm for invasive histology or 3mm for DCIS, TS, or T1 disease. Patients receiving neoadjuvant chemotherapy or having multifocal cancer, pure invasive lobular histology, surgical margins < 2mm, a lumpectomy cavity within 5mm of the body contour, or unclear cavity delineation on the planning scan were excluded. Additionally, patients with evaluation PTV (PTV_Eval) volumes exceeding 124cc were excluded based on fat necrosis observed by Timmerman et al. above this threshold [8]. 23 patients were not included in the RAD 1802 study, but were simulated, contoured, and planned with the same methods and intent. For all patients, an isotropic 1cm gross tumor volume (GTV) expansion was utilized for clinical target volume (CTV) generation and an isotropic 3mm CTV expansion was utilized for PTV generation. PTV_Eval volumes were created by carving out the PTV at anatomical boundaries (i.e., lung, rib, chest wall, and 5mm from the skin). PTV_Eval volume ranged from 28.6cc to 217.9cc, with an average of 85.2cc. Patients were prescribed 3000cGy in five fractions, with an average 98.3% of the PTV_Eval receiving 3000cGy in the original clinical plans. Patient characteristics are summarized in Table 1.

**Table 1.**
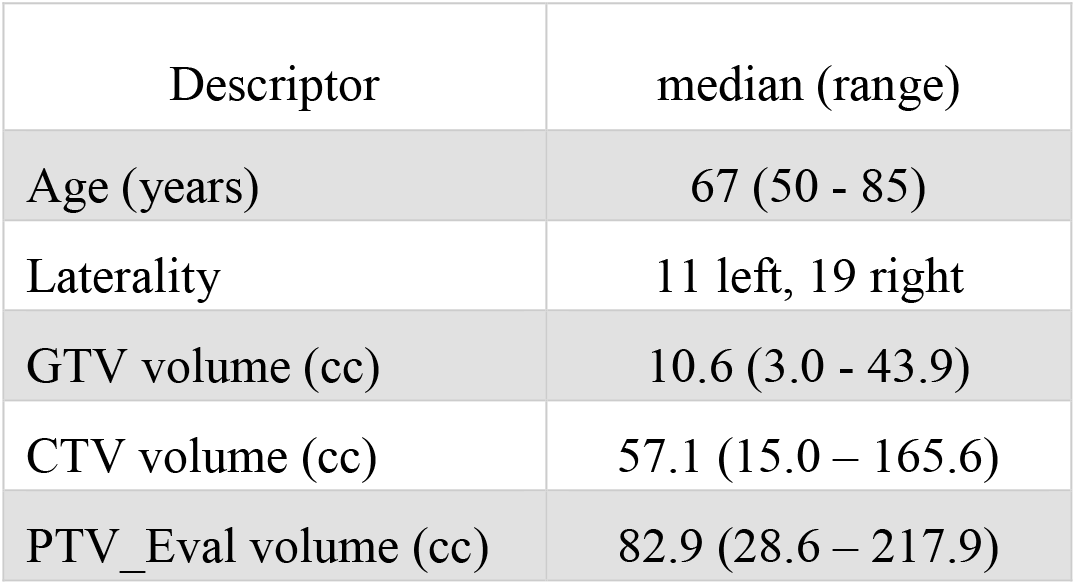
Patient cohort description.

| Descriptor | median (range) |
| --- | --- |
| Age (years) | 67 (50 - 85) |
| Laterality | 11 left, 19 right |
| GTV volume (cc) | 10.6 (3.0 - 43.9) |
| CTV volume (cc) | 57.1 (15.0 – 165.6) |
| PTV_Eval volume (cc) | 82.9 (28.6 – 217.9) |

### 2.2 Treatment Planning

Nine patients previously treated on the Ethos were selected for our tuning cohort (one bilateral patient, four left breast plans, six right breast plans). The tuning cohort was used to establish an Ethos planning template that generated plans meeting RAD 1802 treatment planning goals (Table 2) through iterative planning and fine-tuning of the optimization objectives. A particular emphasis was placed on lowering heart dose to the extent possible while maintaining otherwise similar plan quality to clinical plans. Twenty patients originally receiving supine RT on a Varian TrueBeam Edge were assigned to the validation cohort (seven left breast, thirteen right breast), and were automatically planned using the template resulting from the tuning cohort. Clinically approved Eclipse contours were exported from Eclipse to Ethos and were used for plan generation without modification (i.e., the manually-generated Eclipse lung contour was used in optimization instead of the Ethos auto-contoured lung volume). Ethos validation cohort plans were not reoptimized or renormalized prior to evaluation and were thus evaluated “as-is”.

**Table 2.** APBI planning goals utilized in this study.

| Plan metric | Constraint |
| --- | --- |
| PTV V100% (%) | $\geq 95.0$ |
| Ipsilateral breast V30Gy (%) | $< 20.0$ |
| Ipsilateral breast V15Gy (%) | $< 40.0$ |
| Contralateral breast V5Gy | $< 20.0$ |
| Heart V1.5Gy (%) | $< 5.0$ (right) |
| Ipsilateral lung V9Gy (%) | $< 10.0$ |
| Contralateral lung V1.5Gy | $< 10.0$ |
| Skin D0.01cc (Gy) | $< 39.5$ |
| Rib D0.01cc (Gy) | $< 43.0$ |
| RTOG CI | $< 1.30$ |

Clinical Edge plans were originally calculated with Acuros XB (AXB version 15.5.11, Varian Medical Systems) with heterogeneity correction on and dose-to-water selected. Because Ethos automatically calculates with AXB, dose-to-medium (version 16.1.0), all 20 Edge plans were recalculated using dose-to-medium prior to plan comparison. Recalculations preserved beam geometries and field weightings, but plans were re-normalized to the clinically accepted PTV_Eval prescription isodose coverage. A 2.5mm grid was used for dose calculation in both TPS. The Varian TrueBeam Edge is a stereotactic linear accelerator equipped with a 10MV flattening filter free (FFF) beam, high definition MLCs (HDMLC: 0.25cm in the center, 0.50cm in the periphery), and a maximum dose rate of 2400 MU/min. The Ethos is a CBCT-guided adaptive capable rotational linear accelerator equipped with a 6MV FFF beam, dual stacked and staggered MLC banks as its primary form of collimation, and a maximum dose rate of 800 MU/min [37].

The Ethos pre-defined planning geometries selected for this work include equidistant 9- and 12-field IMRT plans, an ipsilateral 7-field IMRT plan, a 2 full-arc VMAT plan, and a 2 half-arc (180-degree arc span) VMAT plan. While Eclipse optimization is dictated by an internal cost function that varies with assigned priority number, Ethos plans are optimized according to the ascending order of planning objectives submitted in the dose preview workspace. The optimum plan geometry generated from each intent was selected by the reviewing physicist based on adherence to RAD 1802 objectives. Selected Ethos plans were exported to Eclipse, where they were benchmarked dosimetrically against clinically delivered Edge plans. Eclipse and Ethos objective metrics and dose volume histograms (DVH) were extracted via the Eclipse Scripting Application Programming Interface (version 16.1). In addition to presenting the RTOG CI [38], high dose spillage [8] and Paddick gradient index (GI) [39] values were calculated to enable a more holistic plan quality evaluation. The CI, high-dose spillage, and GI are defined in equations

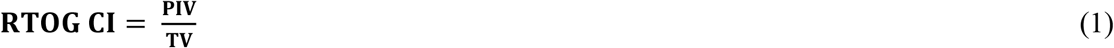

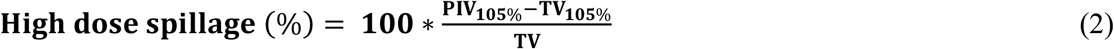

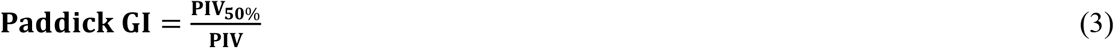

Here PIV and TV are the prescription isodose volume and treated volume (i.e., PTV_Eval volume), respectively. Subscripts specify the isodose volumes evaluated if different than 100%. The Wilcoxon paired, non-parametric test was utilized to test for significant difference between Eclipse and Ethos plan metrics. When conducting multiple tests on the same dependent variable, the likelihood of observing a significant result by pure chance increases. Thus, a Bonferroni correction was applied to adjust for multiple testing, and *p* < 0.004 is considered significant (α = 0.05/12). Statistical analyses were performed in the Python ScyPy library without removal of outliers.

### 2.3 Physician Review

Two board certified radiation oncologists specializing in accelerated partial breast treatment qualitatively evaluated all twenty automatically generated Ethos validation cohort plans according to a previously-utilized in-house grading scheme, which is outlined in Table 3 [40]. To avoid scoring bias, the physicians were not shown the Ethos optimization template before evaluation; in addition, the physicists did not provide feedback or respond to physicians during evaluation, nor were the physicians aware of the cardiac-sparing emphasis of this study. Rather, the physicians graded each plan based on their past clinical experience and their unique interpretation of the scoring criteria. Physicians were not provided case-specific information and performed evaluations solely with anonymous patient identifiers. In cases where plans selected by the physicist would require modification prior to treatment (i.e., a clinically unacceptable physician score of 1-3), the physicians were asked to evaluate their preferred alternative geometry plan for clinical acceptability. The proportion of planning intents that automatically generated at least one plan that the physician deemed clinically acceptable without re-optimization was then evaluated.

**Table 3.** Physician qualitative review grading scheme.

| Score | Description |
| --- | --- |
| 5 | **Use as-is.** Clinically acceptable plan that could be used for treatment without change. |
| 4 | **Minor edits that are unnecessary.** Reviewer prefers stylistic changes but considers current plan acceptable for treatment. |
| 3 | **Minor edits that are necessary.** Reviewer would require changes prior to treatment and the changes, in the judgment of the reviewer, can be implemented by minimal editing of the objectives. |
| 2 | **Major edits.** Reviewer would require changes prior to treatment and the changes in the judgment of the reviewer would require significant modification of the objectives. |
| 1 | **Unusable.** The plan quality is so poor that it is deemed unsafe to deliver, i.e. would likely result in harm to the patient. |

## 3 Results

### 3.1 Planning Template and Intent

From each APBI planning intent submitted in Ethos, five plans with varying geometries were automatically generated. A total of 110 intent and intent revisions were created in this work, equating to the generation and evaluation of 550 unique APBI plans. Ninety intents were required to iteratively plan the nine patient tuning cohort, and the twenty validation cohort patients were each only planned with one unbiased intent. When plans were optimized solely using the RAD 1802 dosimetric objectives in Table 2, many plans failed to meet planning goals. Thus, the planning template in Table 4 was iteratively procured to maximize the likelihood of achieving all planning objectives. The left sided template is shown as an example, but the right sided template is included in supplementary materials. Both templates in XML format are available upon request for easy reproduction of this work by other researchers.

**Table 4.** Ethos left sided APBI planning template. The skin was generated using a 3mm inward expansion of the body surface.

| Priority | Structure | Planning Goal | Acceptable Variation |
| --- | --- | --- | --- |
| 1 | GTV | V3000cGy ≥ 100% | V3000cGy ≥ 99% |
| | GTV | D100% ≥ 3005cGy | D100% ≥ 3000 cGy |
| | CTV | V3000cGy ≥ 99% | V3000cGy ≥ 98% |
| | Heart | V150cGy ≤ 3% | V150cGy ≤ 5% |
| | PTV_Eval | V2850cGy ≥ 99% | V2850cGy ≥ 98% |
| | Heart | Dmean ≤ 150cGy | Dmean ≤ 200cGy |
| | Heart | V700cGy ≤ 0.5% | V700cGy ≤ 10% |
| | Heart | D0.03cc ≤ 1200cGy | D0.03cc ≤ 1500cGy |
| | PTV_Eval | V3000cGy ≥ 97.5% | V3000cGy ≥ 95% |
| | PTV_Eval | D0.03cc ≤ 3700cGy | D0.03cc ≤ 3900cGy |
| | Rib | V3000cGy ≤ 0.80cc | V3000cGy ≤ 1.00cc |
| | _Lung_R | V150cGy ≤ 5% | V150cGy ≤ 10% |
| 2 | _Lung_L | V900cGy ≤ 5% | V900cGy ≤ 10% |
| | _Lung_L | V500cGy ≤ 15% | V500cGy ≤ 20% |
| | _RingInner | V3000cGy ≤ 6% | V3000cGy ≤ 8% |
| | _RingInner | D0.03cc ≤ 3000cGy | D0.03cc ≤ 3000cGy |
| | _RingInner | Dmean ≤ 2000cGy | Dmean ≤ 2200cGy |
| | _Lung_L | V1500cGy ≤ 1% | — |
| | _RingMiddle | D0.03cc ≤ 2000cGy | D0.03cc ≤ 2100cGy |
| | _RingMiddle | Dmean ≤ 1150cGy | Dmean ≤ 2000cGy |
| | _Breast_L - PTV_Eval | V1500cGy ≤ 15% | V1500cGy ≤ 40% |
| | _RingOuter | Dmean ≤ 450cGy | Dmean ≤ 1400cGy |
| | _RingOuter | D0.03cc ≤ 1400cGy | D0.03cc ≤ 1500cGy |
| | _Breast_L - PTV_Eval | V2000cGy ≤ 5% | V2000cGy ≤ 30% |
| | _Breast_L - PTV_Eval | V3000cGy ≤ 2% | V3000cGy ≤ 20% |
| | _Breast_R | V500cGy ≤ 15% | V500cGy ≤ 20% |
| | _Breast_R | V1500cGy ≤ 0.02cc | V1500cGy ≤ 0.03cc |
| | Skin | D0.01cc ≤ 3750cGy | D0.01cc ≤ 3950cGy |
| | Skin | V3650cGy ≤ 8cc | V3650cGy ≤ 10cc |

The template prioritizes GTV coverage the highest, followed by PTV coverage and heart avoidance. The contralateral lung V150cGy was given lower priority in the left-sided than in the right-sided template because heart metrics were more challenging to meet for left sided treatments. This lead the optimizer to spill low dose into the contralateral lung in the absence of a higher priority objective.

The left and right templates were identical besides the contralateral lung V150cGy constraint. The PTV was cropped out of the ipsilateral breast to avoid conflicting objectives prior to optimization (i.e., asking the optimizer to irradiate the PTV but spare the breast + PTV). The entire ipsilateral breast (including PTV) was designated as a report only structure and was thus not optimized. The template contains three rings constituting seven objectives focused solely on conformity, fall-off, and limiting high dose spillage. The inner, middle, and outer rings are derived from (0 - 0.5)cm, (0.5 - 1.0)cm, and (1.0 – 3.0)cm PTV_Eval expansions inside of the Body, respectively.

### 3.2 Plan Selection

The twenty patient validation cohort was originally treated on the Edge using 6-field (n = 1), 8-field (n = 1), and 9-field IMRT (n = 5), as well as 2 partial VMAT arcs (n = 13). Validation cohort plan geometries chosen by the physicist to benchmark against the clinical Edge plans are as follows: seven equidistant 9-field plans (35%), four equidistant 12-field plans (20%), five ipsilateral 7-field plans (25%), three VMAT plans with 2 partial arcs (15%), and one VMAT plan with 2 full-arcs (5%). Because sparing the heart is a primary emphasis of this study, Figure 1 shows Ethos and Eclipse axial, sagittal, and coronal dose distributions (100cGy – 3800cGy) for the manually generated plan with the highest heart V150cGy metric. As is visually evident, the Ethos IOE automatically produces an equidistant 9-field IMRT plan with significant cardiac sparing relative to the manual lateral 6-field IMRT plan.

**Figure 1.**
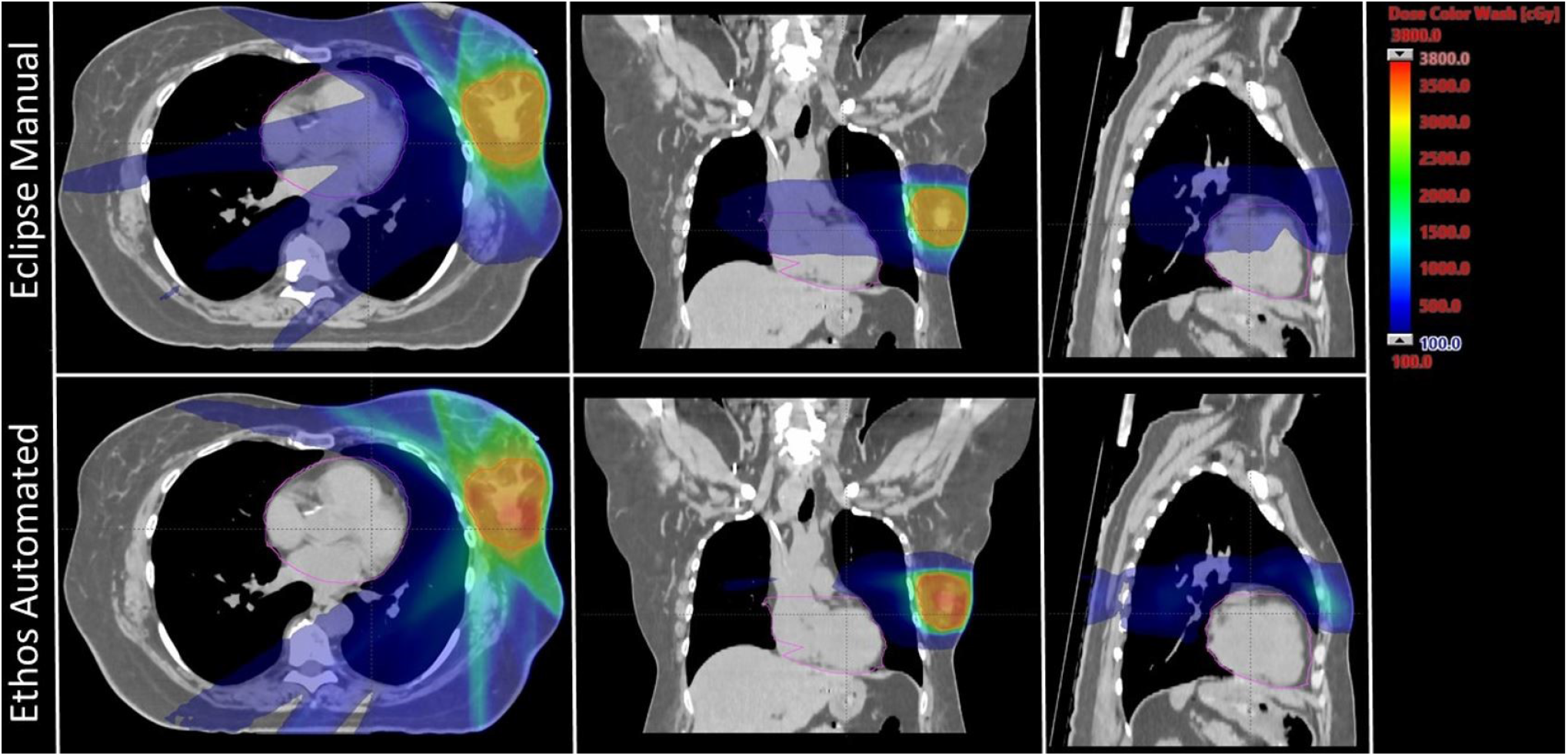
Axial, coronal, and sagittal dose distributions of the manual plan with the highest heart V1500cGy metric for both Eclipse and Ethos. The Eclipse and Ethos plans utilize 6 lateral fields and 9 equidistant fields, respectively. The planning target volume and heart were contoured in red and pink, respectively.

### 3.3 Dosimetry Evaluation

The proposed template automatically generated plans meeting all RAD 1802 objectives for 85% (17/20) of plans without reoptimization. Three initially-selected plans failed to meet the contralateral lung V150cGy constraint. No other constraints were violated in any validation cohort plan. 90% (18/20) of the manually generated clinical Edge plans met all objectives; one plan had less than 95% of the PTV receiving prescription dose and one plan exceeded the contralateral lung V150cGy constraint. Boxplots showing validation cohort metric summaries for Ethos and Eclipse are displayed in Figure 2. Ethos plans had greater PTV_Eval V100% coverage (*p* = 0.01), decreased heart V1500cGy (*p* < 0.001), but increased contralateral breast V500cGy and skin D0.01cc. (*p* = 0.03 and *p* = 0.03 respectively). Although several metrics have medians and interquartile ranges (IQR) that differ, only the heart V150cGy distributions are significantly different when a Bonferroni correction is applied to adjust for multiple hypothesis testing. The Eclipse left sided heart V150cGy IQR and maximum value and are 21.7% and 29.3%, respectively, whereas they are 0.4% and 0.5% for Ethos. The minimum Eclipse right sided heart V150cGy metric is 1.4% while the maximum Ethos V150cGy metric is 0.6%.

**Figure 2.**
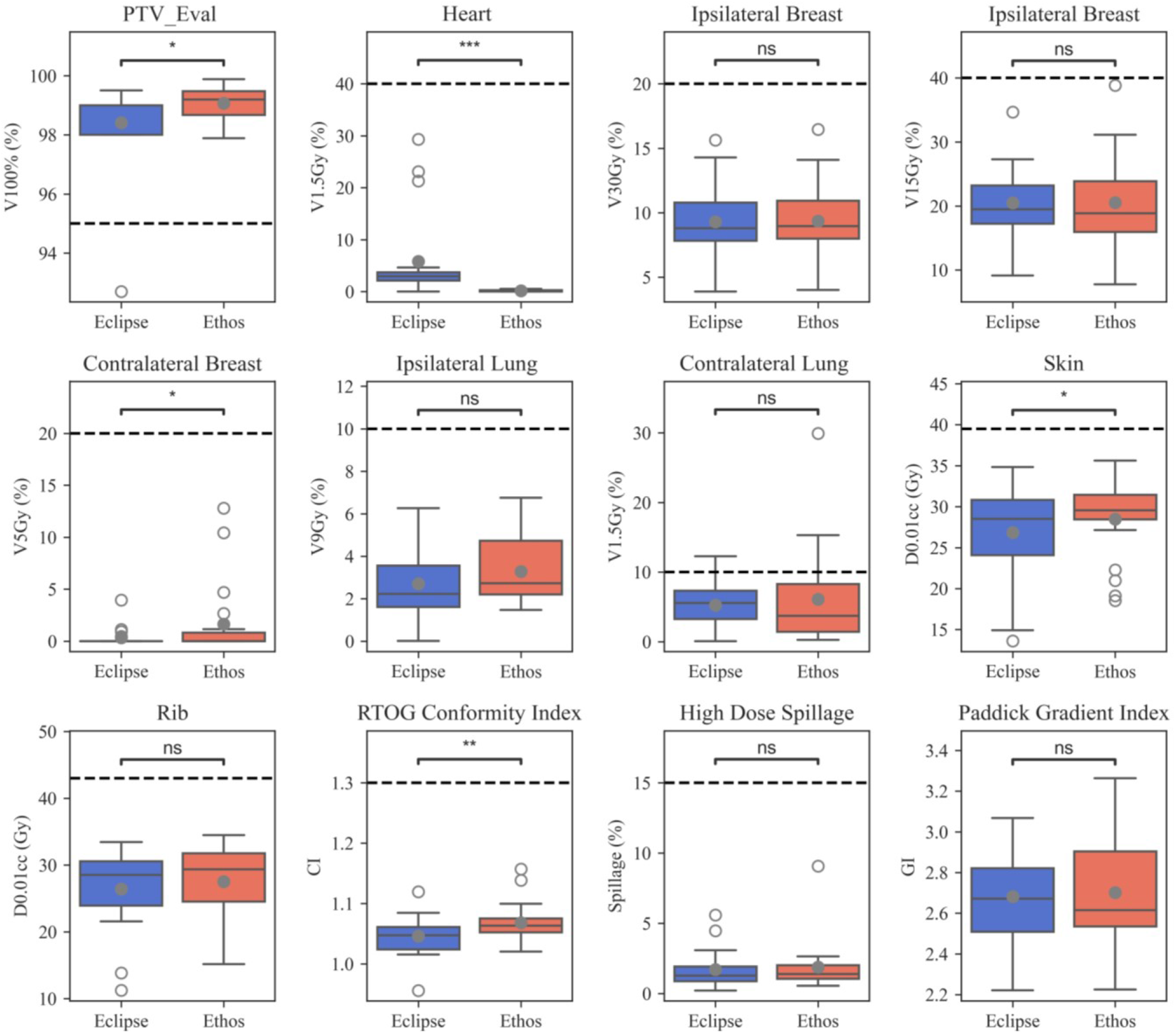
Boxplots summarizing manual Eclipse and Automated Ethos validation cohort planning metrics. Open and closed circles indicate outlier and mean values, respectively. Significance values for the difference between TPS metric distributions were obtained via the Wilcoxon signed rank test and are stratified as follows: ns (not significant): (0.05, *p*, 1.00]; *: (0.01, *p*, 0.05]; **: (0.001, *p*, 0.01]; ***: (0.0001, *p*, 0.001].

All Ethos and Eclipse plans easily met the 1.30 CI planning objective; one Ethos outlier was much greater than all other plans and one Eclipse plan had a CI of 0.95 due to 92.7% PTV_Eval coverage. The median Eclipse and Ethos CI were 1.05 and 1.06, respectively. 100% of the Eclipse and Ethos plans met the 15% high-dose spillage constraint planning suggested in the Timmerman study [8].

There is little discernable difference in high-dose spillage and GI distributions between both TPS when outliers are excluded. Ethos plans generally had more compact high-dose spillage values, but a greater GI IQR. The median Ethos GI was lower, but mean values were similar. While Eclipse CI values were lower than Ethos (*p* = 0.01), there were no significant quality metric differences between both TPS.

Validation cohort mean DVHs with standard deviation bounds are presented for both TPS in Figure 3. The inferior/superior triangle tips illustrate planning objectives and the insets elucidate DVH difference between both TPS (i.e., Ethos volume minus Eclipse volume as a function of dose). Ethos had superior PTV coverage between approximately 2950cGy and 3150 cGy, but a lower portion of the target received above 105% of prescription dose, which is generally preferred for SBRT. Ethos significantly spares the heart above 25cGy, and on average, the heart volume receiving 100cGy was 10.8% less for automated Ethos plans. All left sided Ethos plans were substantially below the right sided planning objective. While the ipsilateral breast DVH curves are similar for high doses, Ethos spares the breast below approximately 1100cGy, with a reduction of 3% breast volume receiving 600cGy. Ethos automated plans had overall higher ipsilateral lung dose above 250cGy, but the discrepancy between plan types was at most 1.4%. The template presented here generated plans with generally inferior contralateral breast dose; 3.4% additional volume received 230cGy. Because the Ethos planning approach heavily spared the heart, automated planning also resulted in much lower contralateral lung dose, with 6.5% less volume receiving 100cGy on average.

**Figure 3.**
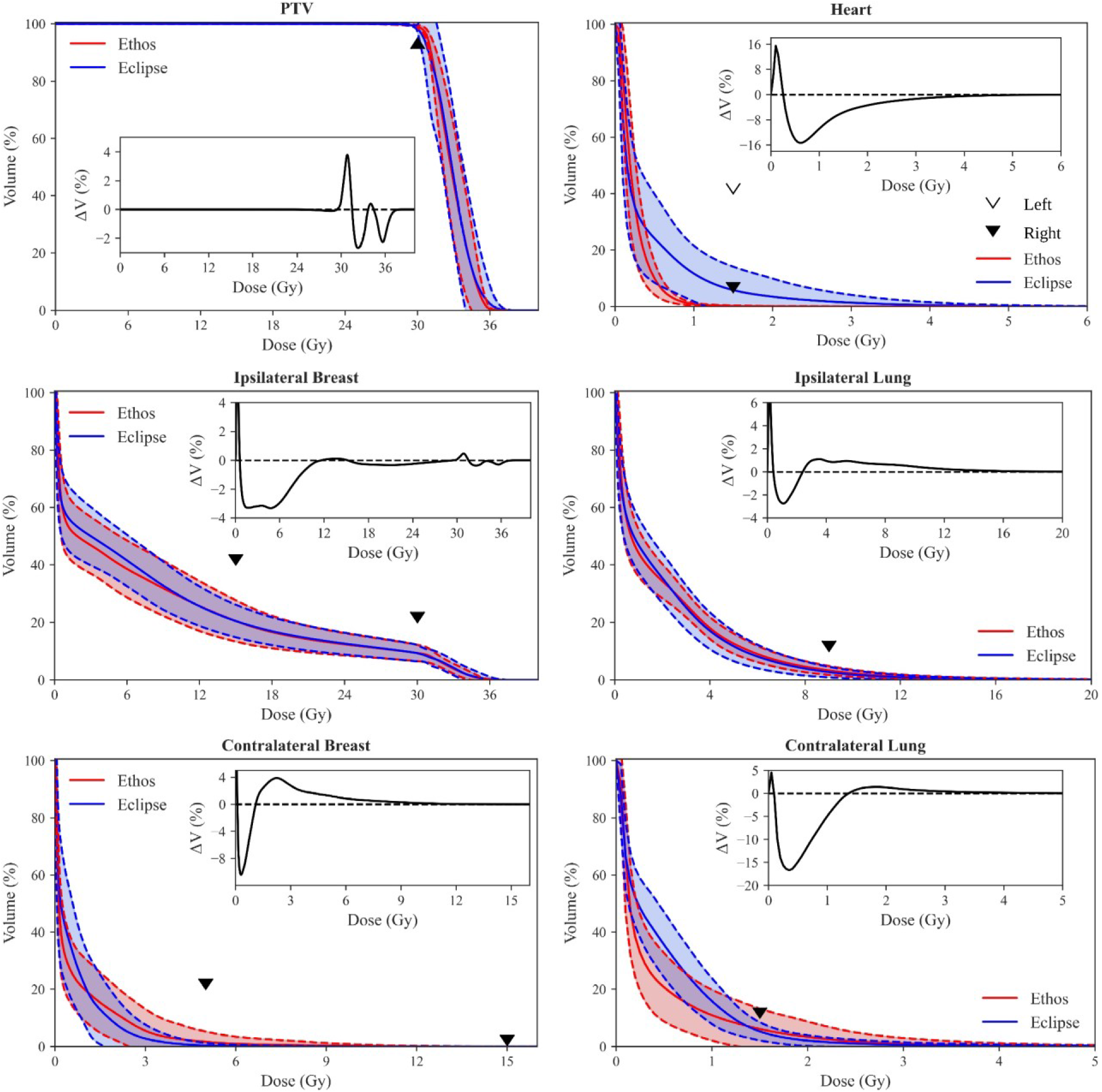
Population DVH comparison of Eclipse manual and Ethos automatic plans. Shaded areas show the mean ± standard deviation of all validation cohort data, and the inferior/superior point of triangles illustrate RAD 1802 planning objectives. Insets show the difference between mean population DVHs (i.e., Ethos mean volume minus Eclipse mean volume). Inset axes were sized for optimal visualization.

### 3.4 Qualitative Evaluation

The physician score summary for physicist-selected Ethos validation cohort plans is shown in Table 5. Physicians A and B considered 75% (15/20) and 90% (18/20) of plans clinically acceptable (scores of 4 or 5) without modification, respectively. 75% of the selected plans (15/20) received a clinically acceptable score from both reviewing physicians. The mode scores of physicians A and B are 4 and 5, respectively. When physicians scored the physicist-selected plan 3 or lower, they then evaluated the alternate plan geometries generated from the same treatment intent and scored the plan they favored most. The five plans receiving a score of 3 from physician A received one 4 and four 5s when alternate plans were evaluated. The two plans receiving a score of 3 from physician B received one 3 and one 4 when alternate plans were evaluated. Thus, at least one plan of treatable quality was automatically generated using the proposed planning approach for 100% of intents for physician A and 95% of intents for physician B.

**Table 5.** Qualitative scoring summary of plans selected by the physicist for physicians A and B.

| Physician | Score |  |  |  |  |
| --- | --- | --- | --- | --- | --- |
|  | 1 | 2 | 3 | 4 | 5 |
| A | 0 | 0 | 5 | 11 | 4 |
| B | 0 | 0 | 2 | 6 | 12 |

Four plans received a score of 3 from physician A due to the lateral extent of 1500cGy streaking prevalent in IMRT plans. One plan received a 3 because physician A preferred the contralateral breast and lung V500cGy be further reduced given favorable patient anatomy. Both plans receiving a score of 3 from physician B were penalized due to lateral extent of 1500cGy streaking. However, physician B further specified that they would have considered whole breast treatment over APBI for the plan receiving a score of 3 even after alternate plan evaluation, primarily due to challenging anatomy and target location.

## 4 Discussion

In this work, we evaluated APBI plans automatically generated from a standard planning approach in the Ethos adaptive workspace; nine patients (ten plans) were iteratively re-planned until desired quality was achieved and twenty validation cohort patients were only planned once using the resulting template. 85% of selected validation cohort plans met all planning objectives with significant reduction in heart dose, and physicians A and B scored 75% and 90% of physicist-selected plans as clinically acceptable, respectively. Physicians A and B deemed at least one automatically generated plan clinically acceptable, without modification, 100% and 95% of the time, respectively.

Four patients in this study with PTV_Eval volumes > 124cc (two in the tuning cohort, two in the validation cohort) received APBI treatment despite failing to meet RAD 1802 inclusion criteria. The treating physician for these cases, who is also the RAD 1802 principal investigator, was comfortable exceeding this threshold due to personal APBI experience, and because these patients had larger breasts or were receiving re-irradiation. Ipsilateral breast V3000cGy and V1500cGy objectives were achieved for all four patients.

Plans initially treated on the Edge were of high-quality, with cardiac dose levels below the UAB RAD 1802 objectives for all 20 validation cohort plans. The maximum Eclipse right and left sided V150cGy metrics are 4.6% and 29.3%, respectively. If one adopts the linear no-threshold model for radiation induced cardiac toxicity, it becomes imperative that the planner continue to minimize heart dose, even below acceptable levels, so long as the net effect on target coverage and sparing of other OARs is not detrimental. To that end, the authors argue that leveraging the Ethos to spare even 100cGy is clinically meaningful, so long as other Ethos plan characteristics are similar in quality to manual Eclipse plans. It should also be mentioned that the OAR dose being spared would be greater were the 600cGy x 5 hypo-fractionated scheme converted to 200cGy equivalent fractions.

Ethos plans were slightly, but consistently, less conformal than Eclipse plans. While some of this discrepancy may be attributed to template design and optimizer differences, it is due at least in part to tertiary collimation width. The double banked, 10mm width Ethos MLC bank is staggered, effectively producing 5mm width MLCs. The Edge has 2.5mm central HDMLC leaves, resulting in twice the collimation resolution. It is reasonable to assume that Ethos plans would see some measurable reduction in CI and high-dose spillage were the MLC width halved. However, Automated Ethos plans had superior CI values (1.07± 0.05) compared to the 30Gy arm published by Timmerman et al. using the Cyberknife (1.22 ± 0.10) [7]. It is also important to note that the mean Ethos validation cohort target volume was smaller than the mean 30Gy arm Cyberknife cohort target volume (Ethos: 77.6cc; Cyberknife: 80.9cc), and CI typically decreases with increasing target size. Thus, the authors argue that the automated plans presented here, while slightly less conformal than Edge plans, are still of high-quality. Further studies are required to deconflate the effects of the different collimators and optimization engine on Ethos plan quality.

The upper Ethos outlier for CI and high dose spillage originated from one plan. This plan presented challenging and abnormal patient anatomy which elucidates a fundamental limitation of this study: fixed beam geometries. The target of interest was the smallest PTV_Eval in the validation cohort and located medially in the upper, inner breast quadrant. The standard field geometries failed to address the patient-specific anatomy; the 2 partial arc and lateral IMRT field geometries span angles from 0° to 180°, clockwise, and the equidistant 9 and 12-field IMRT geometries only space fields every 40° and 30°, respectively. Given the very medial nature of this target, it would have benefitted from partial arcs or densely placed lateral IMRT fields ranging from -90° to 90°. This example highlights that the proposed template does not negate the need for dosimetrist involvement or patient-specific anatomy review; it is expected that abnormal target location or anatomy will require beam geometry modification prior to planning in some instances.

Both reviewing physicians performed a slice-by-slice evaluation of all validation cohort plans. Physicians considered disease extent and location, anatomy favorability, dose distribution shape, and PTV undercoverage in addition to verifying satisfactory DVH metrics. IMRT plan geometries tended to have comparable or even improved GI relative to VMAT, leading the reviewing physicist to select many IMRT plans for further evaluation. However, physician A strongly preferred the consolidated shape of VMAT 1500cGy isodose lines compared to IMRT, which tended to exhibit greater lateral extent but similar volume. Physician B was not as opposed to 1500cGy streaking, except in more serious cases. This highlights the role of personal preference when reviewing plans qualitatively.

While we observed stylistic differences in plan evaluation between the two physician raters, the template provides a mechanism to standardize practices across practitioners, resulting in a large majority of evaluated plans considered acceptable during qualitative review. A future prospective analysis will elucidate if any changes are made after the proposed template is clinically commissioned for use outside of this study.

Artificial intelligence (AI) promises to revolutionize every aspect in radiation oncology care, and has already made a profound impact in enabling the clinical implementation of online adaptive radiotherapies [nnn41; 42]. From automated contouring [43; 44; 45] to radiotherapy dose estimations [46; 47; 48], AI applications are playing a key role increasing efficiency and, often times, improving quality of care through more consistent radiotherapy [49]. For example, studies have shown that auto-contouring can significantly save contouring time, providing the critical time savings needed to minimize patient motion during online adaptive treatment design and delivery [50]. While most clinical applications currently focus on efficiency improvements, we can expect that in the near future clinical teams will be supported by various AI-driven clinical support systems to compliment decision-making during adaptive treatment’s design and delivery. In the current study, we evaluate radiotherapy treatment plans generated using Varian’s IOE, which uses an artificial intelligence driven optimization process to automatically generate radiotherapy treatment plans. Our study shows that this novel optimization engine provides high-quality APBI treatment plans for a large majority of cases (with no planner interaction) after defining a robust planning template through a data-driven iterative approach.

APBI treatments were transitioned from the Edge to the Ethos in 2021 at our institution, and APBI treatment for 17 patients has been successfully completed in the Ethos adaptive workspace. During the first course of adaptive treatment on the Ethos, we noticed that the GTV location, volume, and shape changed from simulation to first fraction, and between each subsequent fraction. Consequently, adapted plans significantly spared OARs compared to scheduled plans (i.e., initial treatment-approved plans recalculated onto daily CBCT anatomy). Therefore, even though automated Ethos plans are overall similar in quality to manual Eclipse plans, the added benefit of daily CBCT based adaption vastly outweighs whatever slight deficiencies might exist in the proposed Ethos planning approach (i.e., higher Ethos contralateral breast dose). The impact of daily adaptation on both plan quality and patient outcomes warrants further investigation. Other future projects include implementing the APBI template presented here into our clinical workflow and continuing to generate planning templates for other sites.

The manuscript presented here, including study design and analysis, was developed for consistency with recently published RATING guidelines for generating high-quality planning studies (RAdiotherapy Treatment plannINg study Guidelines) [51]. The authors’ self-assessment score was 94% (195/207) and the accompanying grading template is added to the supplemental materials.

Although APBI planning is challenging due to proximity of many OARs and the need for conformity and steep dose gradients, the Ethos templates investigated in this work automatically generate high-quality left- or right-sided APBI plans. Ethos plans had similar target coverage, reduced heart dose, and otherwise similar OAR dose to manual Eclipse plans. 85% of validation cohort plans met all planning objectives, and only the contralateral lung V150cGy objective was violated for any plan.

Physicians A and B scored at least one plan from each intent of clinically acceptable quality, without reoptimization, 100% and 95% of the time, respectively. Therefore, the approach summarized here enables consistent and high-quality generation of Ethos APBI plans with a specific emphasis on minimizing heart dose.

## Supporting information

RATING score

## Data Availability

The raw data supporting the conclusions of this article will be made available by the authors, without undue reservation.

