## Supplemental table for "Leveraging intelligent optimization for automated, cardiac-sparing accelerated partial breast treatment planning"

Supplementary Table 1 | Ethos right sided APBI planning template. The skin was generated using a 3mm inward expansion of the body.

| Priority | Structure | Planning Goal | Acceptable Variation |
| --- | --- | --- | --- |
| 1 | GTV | V3000cGy ≥ 100% | V3000cGy ≥ 99% |
|  | GTV | D100% ≥ 3005cGy | D100% ≥ 3000 cGy |
|  | CTV | V3000cGy ≥ 99% | V3000cGy ≥ 98% |
|  | Heart | V150cGy ≤ 3% | V150cGy ≤ 5% |
|  | PTV_Eval | V2850cGy ≥ 99% | V2850cGy ≥ 98% |
|  | Heart | Dmean ≤ 150cGy | Dmean ≤ 200cGy |
|  | Heart | V700cGy ≤ 0.5% | V700cGy ≤ 10% |
|  | Heart | D0.03cc ≤ 1200cGy | D0.03cc ≤ 1500cGy |
|  | PTV_Eval | V3000cGy ≥ 97.5% | V3000cGy ≥ 95% |
|  | PTV_Eval | D0.03cc ≤ 3700cGy | D0.03cc ≤ 3900cGy |
|  | Rib | V3000cGy ≤ 0.80cc | V3000cGy ≤ 1.00cc |
| 2 | _Lung_R | V900cGy ≤ 5% | V900cGy ≤ 10% |
|  | _Lung_R | V500cGy ≤ 15% | V500cGy ≤ 20% |
|  | _RingInner | V3000cGy ≤ 6% | V3000cGy ≤ 8% |
|  | _RingInner | D0.03cc ≤ 3000cGy | D0.03cc ≤ 3000cGy |
|  | _RingInner | Dmean ≤ 2000cGy | Dmean ≤ 2200cGy |
|  | _Lung_L | V150cGy ≤ 5% | V150cGy ≤ 10% |
|  | _Lung_R | V1500cGy ≤ 1% | _ |
|  | _RingMiddle | D0.03cc ≤ 2000cGy | D0.03cc ≤ 2100cGy |
|  | _RingMiddle | Dmean ≤ 1150cGy | Dmean ≤ 2000cGy |
|  | _Breast_R - PTV_Eval | V1500cGy ≤ 15% | V1500cGy ≤ 40% |
|  | _RingOuter | Dmean ≤ 450cGy | Dmean ≤ 1400cGy |
|  | _RingOuter | D0.03cc ≤ 1400cGy | D0.03cc ≤ 1500cGy |
|  | _Breast_R - PTV_Eval | V2000cGy ≤ 5% | V2000cGy ≤ 30% |
|  | _Breast_R - PTV_Eval | V3000cGy ≤ 2% | V3000cGy ≤ 20% |
|  | _Breast_L | V500cGy ≤ 15% | V500cGy ≤ 20% |
|  | _Breast_L | V1500cGy ≤ 0.02cc | V1500cGy ≤ 0.03cc |
|  | Skin | D0.01cc ≤ 3750cGy | D0.01cc ≤ 3950cGy |
|  | Skin | V3650cGy ≤ 8cc | V3650cGy ≤ 10cc |
